# Modelling of the second (and subsequent) waves of the coronavirus epidemic. Spain and Germany as case studies

**DOI:** 10.1101/2020.06.12.20129429

**Authors:** Francisco de Castro

## Abstract

The first wave of the coronavirus pandemic is waning in many countries. Some of them are starting to lift the confinement measures adopted to control it, but there is considerable uncertainty about if it is too soon and it may cause a second wave of the epidemic. To explore this issue, I fitted a SEIR model with time-dependent transmission and mortality rates to data from Spain and Germany as contrasting case studies. The model reached an excellent fit to the data. I then simulated the post-confinement epidemic under several scenarios. The model shows that (in the absence of a vaccine) a second wave is likely inevitable and will arrive soon, and that a strategy of adaptive confinement may be effective to control it. The model also shows that just a few days delay in starting the confinement may have caused and excess of thousands of deaths in Spain.

## Introduction

The epidemic of coronavirus (SARS-CoV-2) has spread all over the world since January 2020. Most governments have taken measures in trying to limit its spread and to reduce its severity, with greater or lesser success. The most common, non-pharmaceutical measures are directed to reduce the rate of contacts between people, thus reducing the transmission rate. Among other possibilities, these measures may entail forbidding large assemblies of people, closing of schools and universities, closing of businesses, home isolation of confirmed or suspected cases and tracing their contacts, etc. In what follows I refer to all these measures collectively as confinement.

The rigor of these measures and their timing vary among countries, or even between regions. Some countries adopted measures very early in their epidemic while others took more time. At present (June 2020) in many countries the epidemic has passed its peak and is quickly decreasing, which is leading many governments to start lifting the confinement. Although this is done with a phased, gradual approach, there is still considerable uncertainty about the consequences, and if it may cause a resurgence of the virus and a second wave of cases.

It is difficult to know exactly the effect that the end of confinement will have on the transmission rate and, therefore, on the number of infections. For this reason, predictive models fitted to specific situations are useful as tools to predict the probability of a second (and subsequent) waves, their potential amplitude and to test strategies for their control before they arrive.

Here, I fitted a SEIR model with time-dependent transmission and mortality rates to data from Spain and Germany as case studies. With the fitted model, I simulated the epidemic in both countries post-confinement to show the possibility of a second wave and its potential amplitude and timing under different scenarios. Additionally, I used the model to explore the option of adaptive confinement to control the second and subsequent waves.

## Methods

### All the calculations were done with MATLAB (2018)

### SEIR model

I used a standard SEIR model modified with time-dependent transmission and mortality rates. SEIR models classify the population into four classes: susceptible (*S*), exposed (*E*, infected but not yet infectious), infected (and infectious, *I*), and recovered (*R*). The total population is *N* = *S*+*E*+*I*+*R*. The model is represented by the equations:

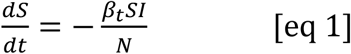

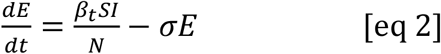

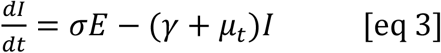

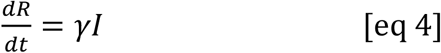

where: *β*_*t*_ is time-dependent transmission rate (eq. 7), *σ* is the loss of latency rate, *γ* is recovery rate and *μ*_*t*_ is time-dependent, disease-induced mortality rate (eq. 8). This formulation assumes a complete mixing of the population and no population dynamics (i.e. births and natural mortality are considered negligible within the time frame of the epidemic). It also assumes that surviving the disease confers permanent immunity.

Two additional equations are included to keep track of the accumulated number of deaths (*D*) and accumulated infections (denoted *AI*, to avoid confusion with currently infected *I*), which are later used to fit the model to data. These are:

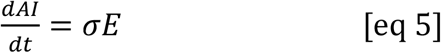

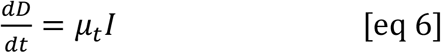

Transmission rate *β*_*t*_ and mortality rate *μ*_*t*_ are not constants but a function of time, to capture the effect of measures taken to control the epidemic. Regarding transmission, this refers to the confinement of a large part of the population and the quarantine of detected cases. Regarding mortality rate, the time-dependency was introduced because a variable mortality rate achieved a better fit, and, with constant mortality, the deviation between model and data suggested a decrease of mortality rate after confinement measures were put in place. Time-dependent transmission and mortality rates are modelled with sigmoid functions, as:

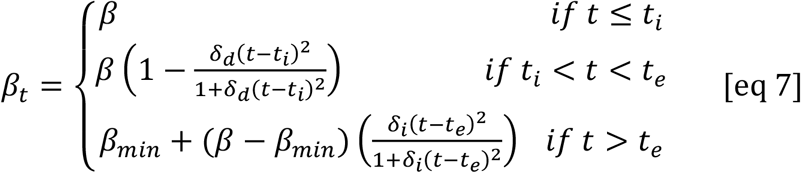

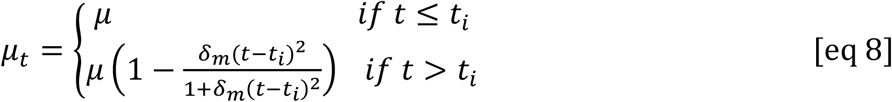

where *β* is the initial transmission rate, without any measures to limit contact between people, and *μ* is the same for the mortality rate. *t* is time (days) from the first detected case, and *t*_*ι*_ is the initial day of confinement minus 2, and *t*_*e*_ is the end of confinement. *δ*_*d*_ is the rate of *decrease* of transmission because of the control measures, while *δ*_*m*_ is the same for mortality. *β*_*min*_ is the value of *β*_*t*_ at time *t*_*e*_, and *δ*_*i*_ is the rate of *increase* of transmission after *t*_*e*_ (see fig. 1 for an example of *β*_*t*_).

**Fig 1.**
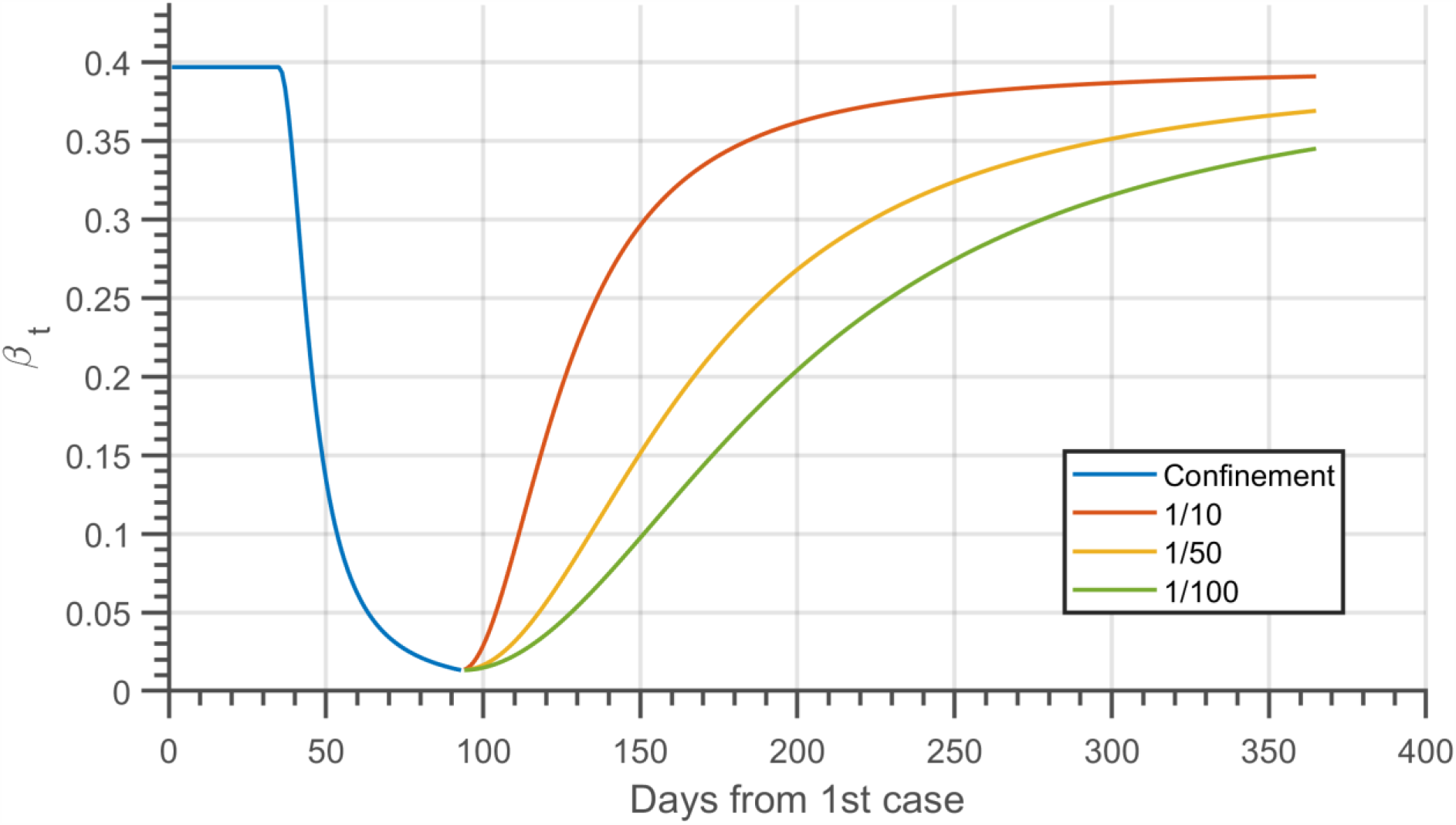
Evolution of time-dependent transmission rate (*β*_*t*_) for Spain before, during, and after confinement. The blue line shows the initial phase with constant *β* (up to day 40) and the decline of transmission with confinement, until day 93. The rate of decrease (*δ*_*d*_) is estimated from model fitting. After day 93 each line represents a different speed of confinement release, as a fraction of the speed of decrease. Red: 1/10, Orange: 1/50 and Green: 1/100.

### Fitting of the model

To fit the model, I used data on daily cases and daily deaths from the European Centre for Disease Control (ECDC 2020) and data of recovered from the John Hopkins University github (CSSE 2020), because the ECDC does not include recovery data. Accumulated time series were generated from daily values. Only values up to the end of the confinement period were used in the fitting, so only the first two lines of eq. 7 are pertinent in this case. At the time of writing there is not yet enough data after confinement to find a good fit for parameter *δ*_*i*_.

The date of start of confinement is not always easy to define, as measures may have been taken incrementally along several days or weeks or may have been adopted at different times in different regions of a country. I relied on published news about the confinement to choose *t*_*i*_ and *t*_*e*_ for Spain and Germany. For Spain: *t*_*i*_ was the 14^th^ of March and *t*_*e*_ the 10^th^ of April. For Germany *t*_*i*_ was the 23^rd^ of March and *t*_*e*_ the 20^th^ of April.

When comparing the model to data we must consider the question of detectability. Most of the cases are undetected, either because they are mild or even asymptomatic or because they are not tested for the virus for different reasons. In the model all cases are known exactly, so to make a meaningful comparison with the data the model values were multiplied by a probability of detection, *p*_*det*_. Recent extensive serological studies in Spain have shown a prevalence of 5%, or around 2.3 million people (Instituto de Salud Carlos III, 2020). This is approximately ten times the number of diagnosed cases in Spain, which suggests a value of 0.1 for *p*_*det*_. I used the same value for Germany. Deaths, on the other hand, are assumed to be known exactly. If more deaths were due to the epidemic, the parameter estimates could change, particularly mortality rate, but until a more accurate estimation of deaths is available, I rely on data as published.

The fitting is done by least squares, combining four time-series: accumulated infected, accumulated deaths, accumulated recovered, and currently infected. The quantity to be minimized is, then:

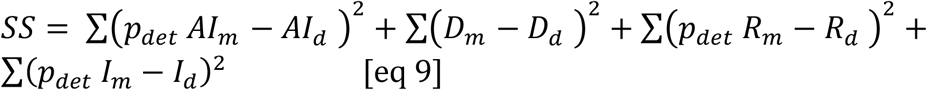

The minimization was done with MATLAB function *lsqcurvefit*. In eq. 9 the subscript *d* stands for data, while subscript *m* stands for model. *AI* is accumulated infected (eq. 5), *D* is accumulated deaths (eq. 6), *R* is accumulated recovered (eq. 4) and *I* are currently infected (eq. 3). The number of current infections is typically not reported, so I calculated it as:

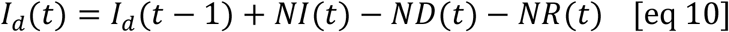

where: *I*_*d*_(*t*) are observed current infections on day *t, I*_*d*_(*t-1*) are the current infections the day before and *NI*(*t*), *ND*(*t*), and *NR*(*t*) are the new infections, new deaths, and new recoveries on day *t*, respectively.

To generate a model prediction with a specified set of parameters the model equations are integrated using MATLAB function *ode45*. This solver uses an explicit Runge-Kutta formula to do the integration. The initial values for each population class were: as susceptible, the total population of the country (World Bank 2019), no individuals exposed, one individual infected, no dead, and no recovered. The initial values for the parameters and their bounds during fitting are shown in Table 1.

**Table 1.** Initial values and bounds of the free parameters of the model. The values of *σ* and *γ* are expressed as the inverse of the latency period and infective period, respectively. The value of *β* is calculated from *γ* and *μ* assuming an *R*_0_ of 3.

| Parameter | Initial value | Lower bound | Upper bound |
| --- | --- | --- | --- |
| $\beta$ | $3 (\gamma + \mu)$ | 0 | 5 |
| $\sigma$ | 1/3.5 | 1/10 | 1 |
| $\gamma$ | 1/15 | 1/60 | 1/4 |
| $\mu$ | 0.01 | 0 | 0.25 |
| $\delta_d$ | $5 \cdot 10^{-4}$ | 0 | 0.25 |
| $\delta_m$ | $5 \cdot 10^{-4}$ | 0 | 0.25 |

After fitting the model, the basic reproductive number (*R*_0_) is estimated (Diekmann et al. 1990, Blackwood & Childs 2018) as:

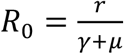

where *r* is the exponential rate of increase of cases at the beginning of the epidemic, which, in turn, is estimated by fitting an exponential function using only the first 50 days of data.

Once a best-fitting model is identified, confidence intervals for all the parameters (and *R*_0_) were estimated by bootstrap, assuming a Poisson error structure and using 1000 realizations (following Chowell 2017). I fitted the model to data from two countries: Spain and Germany because they have shown contrasting epidemic trajectories, which can shed light on the benefits or pitfalls of different control strategies. The parameter estimates and their confidence intervals for both countries are shown in Table 2.

**Table 2.** Parameter estimates and 95% confidence intervals for Spain and Germany. For a description of the parameters, see methods. *R*_0_, while not a model parameter, is shown for its indicative value.

| Parameter | Spain |  |  | Germany |  |  |
| --- | --- | --- | --- | --- | --- | --- |
|  | Estimate | 95% CI |  | Estimate | 95% CI |  |
| $\beta$ | 0.6101 | [0.6023 | 0.6179] | 1.0779 | [1.0736 | 1.0819] |
| $\sigma$ | 0.2578 | [0.2521 | 0.2637] | 0.0863 | [0.0859 | 0.0867] |
| $\gamma$ | 0.0334 | [0.0333 | 0.0335] | 0.0711 | [0.0707 | 0.0715] |
| $\mu$ | $15 \cdot 10^{-4}$ | [$14 \cdot 10^{-4}$ | $16 \cdot 10^{-4}$] | $3.74 \cdot 10^{-4}$ | [$3.63 \cdot 10^{-4}$ | $4.14 \cdot 10^{-4}$] |
| $\delta_d$ | 0.0172 | [0.0169 | 0.0175] | 0.0717 | [0.0709 | 0.0725] |
| $\delta_m$ | 0.0016 | [0.00152 | 0.00176] | $1.17 \cdot 10^{-8}$ | [$2.36 \cdot 10^{-14}$ | $9.78 \cdot 10^{-5}$] |
| $R_0$ | 6.27 | [6.25 | 6.30] | 3.16 | [3.13 | 3.19] |

## Results and simulations

The model reached a very good fit for both countries and for all the components of the population: total infections, current infections, deaths, and recoveries (fig. 2). This good fit lends confidence in using the model as a predictive tool and for the testing of hypothesis or the consequences of control strategies. I used the model to simulate the evolution of the epidemic post-confinement in several scenarios, and the feasibility of an adaptive confinement strategy.

**Fig 2.**
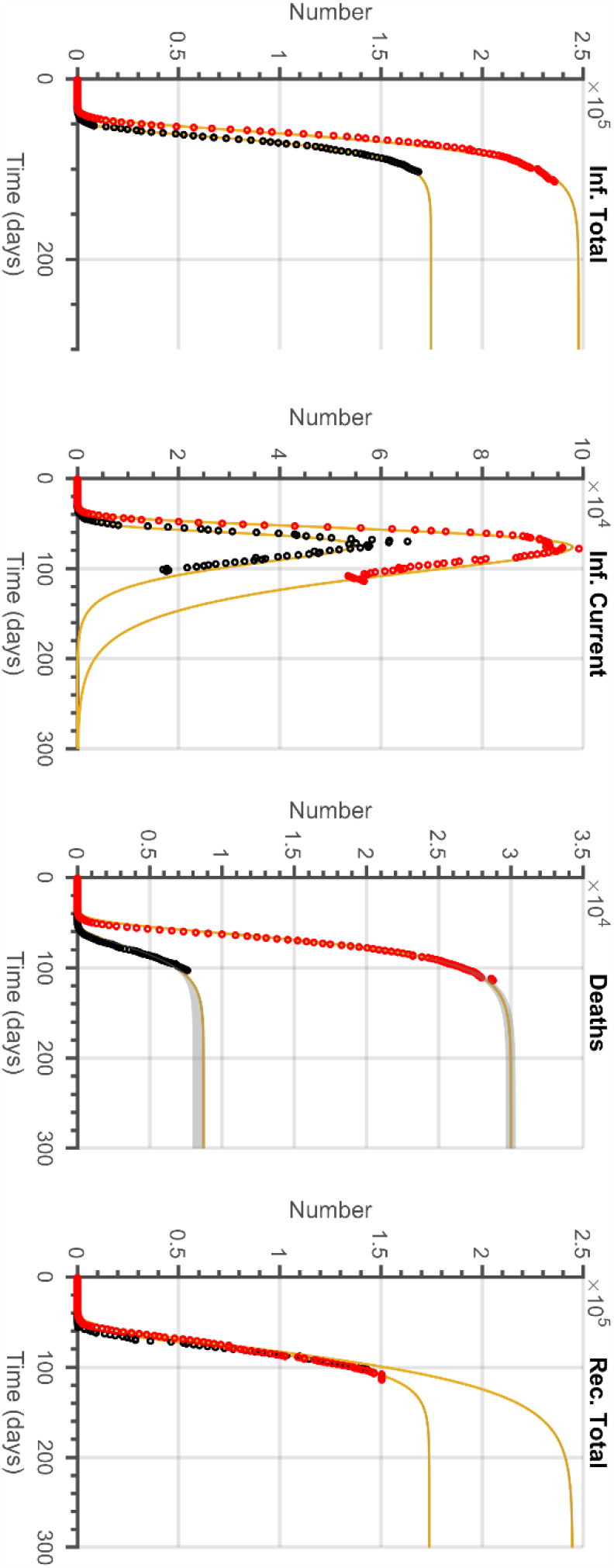
Fit and forecast of the model for Spain and Germany. From left to right the panels represent: accumulated infected, currently infected, deaths, and recovered. Red dots represent Spain and black dots represent Germany. The yellow line is the best-fitting model, forecasted 300 days after the first infection (in each country). Grey areas are 95% confidence intervals for the prediction from a bootstrap procedure (see methods). Confidence intervals are not visible for the most part because they are very narrow.

### The second wave

Many countries, including Spain and Germany, are already lifting the confinement measures, typically in a slow, phased approach. However, there is considerable unease and uncertainty about if the lifting of confinement will provoke a resurgence of the virus and a second wave of cases, and its potential magnitude. It is very difficult to know with any exactitude the effect that the lifting of each measure will have on the transmission rate and, therefore, on the number of infections. However, it seems safe to assume that any increase of transmission rate will be slower than the decrease that happened as a consequence of the confinement. Or, in terms of the model, that δ i ≪ δ d. I used the Spain and Germany models to simulate the possible evolution of the epidemic post-confinement in three scenarios, each characterized by a value of the rate of increase of transmission *δ*_*i*_. The three values are expressed as a fraction of the rate of *decrease* of transmission *δ*_*d*_. Specifically I used: *δ*_*i*_= *δ*_*d*_/10, *δ*_*i*_= *δ*_*d*_/50, and *δ*_*i*_=*δ*_*d*_/100 (fig. 3). It is important to keep in mind that these simulations assume that no further confinement measures would be taken in any case.

**Fig 3.**
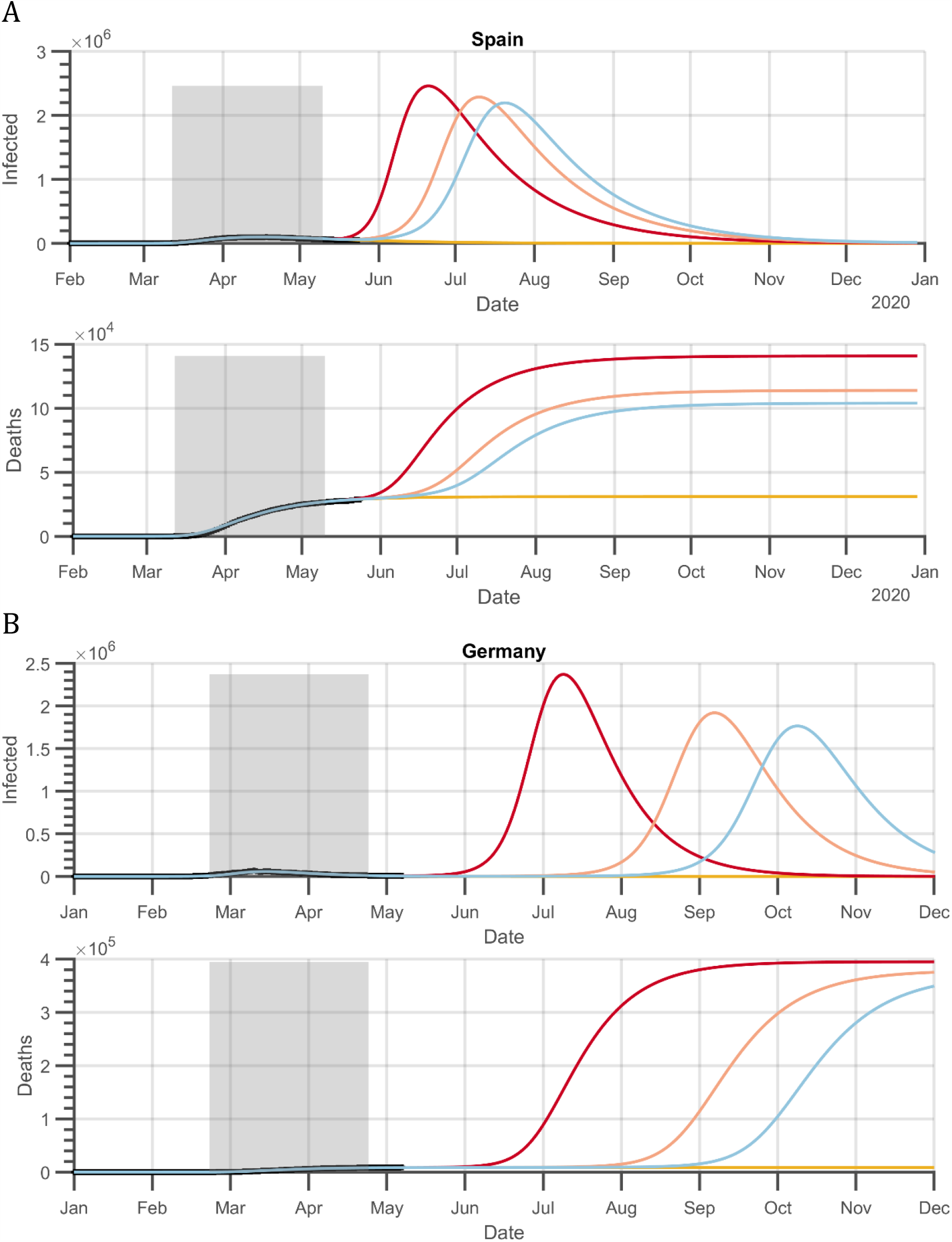
Simulation of the easement of confinement. Panel A shows results for Spain and panel B for Germany. The grey area is the period of confinement. In each panel, the top figure shows current infections and the lower one number of deaths. Each line represents a different speed of confinement release, expressed as a fraction of the speed of *decrease* of transmission during confinement (*δ*_*t*_). Red line: 1/10, pink line: 1/50, blue line: 1/100. The yellow line shows model prediction maintaining confinement.

The results suggest that there will be a second wave in both countries and that their magnitude would dwarf the current wave. In fact, they would be as large as the first wave would have been if no measures at all had been taken. At the peak of the second wave the number of current infections would be between 2 and 3 million and the number of deaths would be in the hundreds of thousands, for both Spain and Germany. The timing of the second wave, on the other hand, would be different in both countries. Spain, in the worst-case scenario, would have the peak of the second wave in mid-June, and in the best-case scenario in late July. Germany, on the other hand, would have a second-wave peak between early July and early October, depending on the scenario.

### Adaptive confinement

The results of the simulation of the second wave strongly suggest that, in the absence of a vaccine or an extremely efficient track-and-trace system, further periods of confinement will be necessary in the near future. I used the Spain-fitted model to simulate a situation in which the confinement is triggered when the number of currently infected exceeds a given threshold, and it is gradually lifted when the number drops back below it (Ferguson et al. 2020). I used the number of currently infected as trigger variable and simulated two scenarios, with a threshold of 3500 and 1000, respectively. The former is the number of infected there were in Spain two days before the “estate of alert” (i.e. confinement) was declared, on the 14^th^ of March. The later was reached approximately four days before, on the 8^th^ of March (fig. 4).

**Fig 4.**
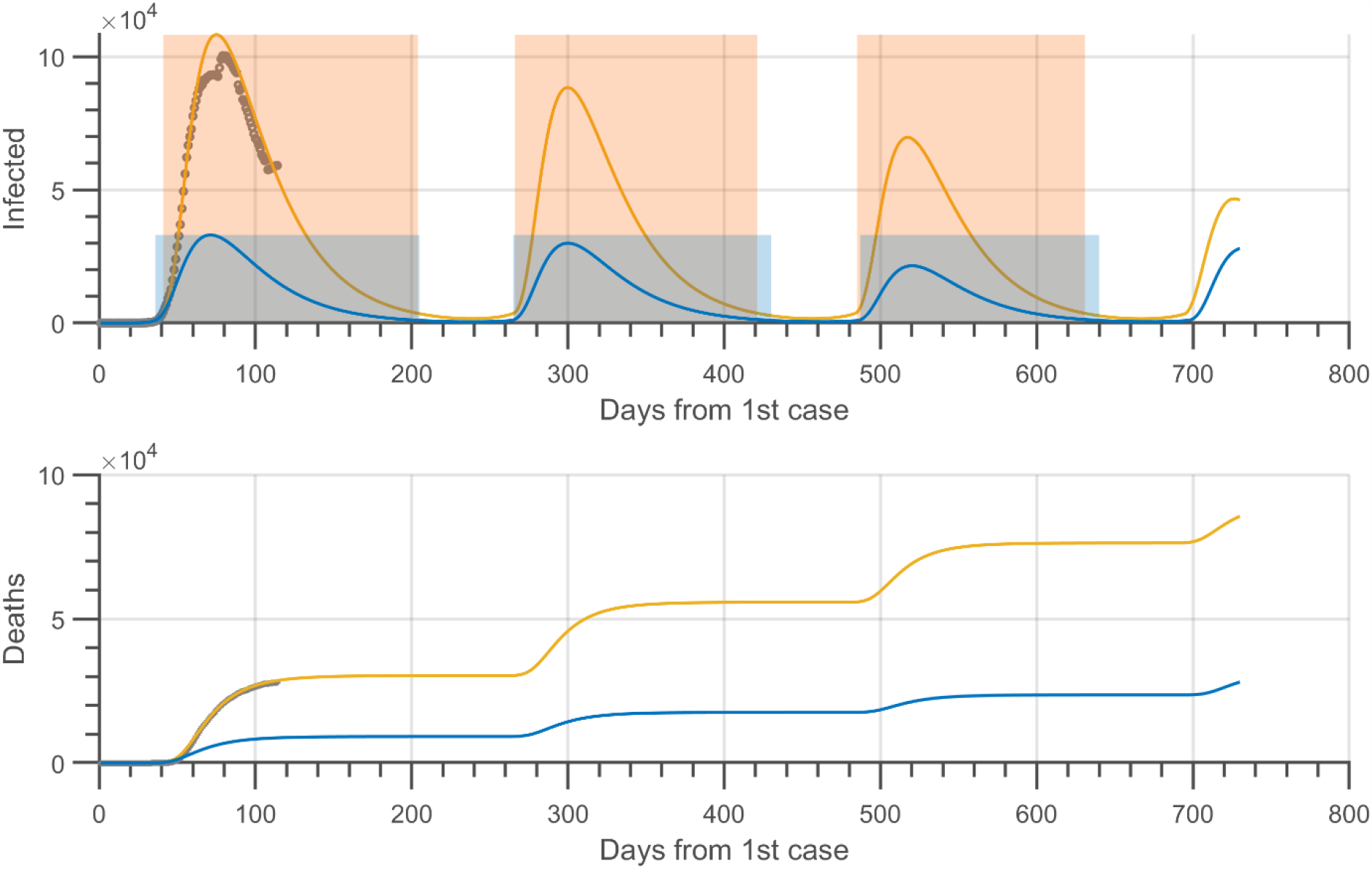
Simulation of adaptive confinement with the Spain-fitted model. The confinement is triggered when the number of active infections exceeds a specified threshold and is *gradually* lifted when the number drops back below it. The threshold was 3500 for the yellow line and 1000 for the blue line. The top panel shows active infections and the lower panel shows deaths. The semi-transparent areas in the top panel mark the periods of confinement (above threshold), in orange for the higher threshold and blue for the lower one.

The results show that the epidemic can be controlled by using this type of adaptive strategy, imposing or easing a lockdown depending on the state of the epidemic. This adaptive model shows three waves of decreasing amplitude (including the present one) in the first two years, with the thresholds mentioned above. Crucially, the simulations also show large differences in the number of infections and deaths depending on the threshold used to trigger the confinement. This fact, given the exponential growth of the number of cases at the start of the epidemic, stresses the importance of taking measures early in the epidemic.

## Discussion

I fitted a SEIR model with time-variable transmission and mortality rates to the COVID19 epidemic data of Spain and Germany, as case studies. The model fitted the data extremely well in both cases, which allows to use it with confidence to predict the future evolution of the epidemic or the outcomes of different control strategies. The fact that the transmission rate, particularly, is not constant but time-dependent allows the model to respond to the confinement of part of the population, which is the most commonly used strategy to control or reduce the epidemic.

The timing of control measures plays an essential role in the model, but it is not straightforward to define this timing in a way that matches reality. In the model, the confinement starts in a specific day for the whole population, but this is typically not the case. In some countries, control measures were taken incrementally as the epidemic progressed, not all in a single day. Furthermore, some measures may be applied at different times in different regions of a country, like it happened in Italy, which does not fit the assumption of homogenous mixing implicit in the model. However, despite this difficulty, the model retains its capacity to fit the observations quite well, thus making it a good tool for prediction and testing.

The estimates of the model parameters show differences between Spain and Germany which could be related to their management of the epidemic (Table 2). Given that Germany’s population is almost twice that of Spain, a stronger epidemic would have been expected there. However, the total cases, deaths and peak of infections were considerably lower. Although the initial transmission rate (*β*) was higher in Germany, the decrease of transmission with confinement was also much faster, and this, coupled with a higher recovery rate and lower mortality, led to a much-restricted epidemic. The basic reproductive number (*R*_*0*_) also offers clues on the severity of the epidemic. It was twice as high in Spain, and higher in both countries than most estimates from other studies (Liu et al. 2020), but still within the range estimated by other authors (Shen et al. 2020, Sanche el at. 2020). Further, by the end of confinement the case fatality rate was 0.5% in Germany and 1.22% in Spain; while the fatalities per hundred thousand people were 11.2 and 64.1, respectively. It would be difficult to attribute any of these differences to specific circumstances. Undoubtedly, they are the combined outcome of multiple factors, but taken together suggest that the management of the epidemic was more effective in Germany.

The simulations of the post-confinement period clearly show that a second wave is probably inevitable, no matter how gradually or carefully the confinement is lifted. The reason is the large pool of infected (mostly undetected) still present among the population when the lifting of confinement measures is started. The model estimate of the number of infected at the end of the confinement was 775×10^3^ in Spain, and 149×10^3^ in Germany. This is undoubtedly much larger than the initial, imported cases that started the epidemic in either country, which makes a second wave very likely and with the potential of being larger than the first. The difference between the two countries in number of infected when confinement was lifted is also the reason for the differences in the timing of the second-wave peak, which the model predicts will happen much earlier in Spain. In the best-case scenario this difference is approximately 2.5 months. To delay a second wave as much as possible in Spain it would be necessary to prolong the confinement measures until the pool of infected has reduced considerably below its present level. However, it would be probably unfeasible to prolong it until the complete disappearance of the disease. The model predicts that the last case in Spain (maintaining confinement) would occur in late November and in Germany by the end of August. A good track-and-trace system may help to reduce the amplitude of the second wave, but it would require a very high efficacy.

Given that a second wave is likely inevitable and that is not possible to maintain the confinement measures until the disease disappears completely, other strategies will be required, like the adaptive policy suggested by Fergusson et al. (2020). The simulations of this strategy with the Spain-fitted model (fig. 4) show that it is feasible to control the epidemic by alternating confinement periods with others of non-confinement. There is a trade-off to consider, however, in using this strategy; setting the threshold higher makes the periods of confinement slightly shorter, but the epidemic waves will be larger and, consequently, there will be more fatalities. Another factor to consider is what to choose as indicator. I used here the number of active infections, while Fergusson et al. (2020) suggested the case incidence in ICU patients. This question is open to debate, but the best indicator (which may be different in each country) would be one that can be measured as accurately and with as little delay as possible.

The model predicts that there would be three waves of diminishing amplitude in two years, including the current one, with the thresholds chosen. The reason for the decrease in amplitude is the increasing percentage of the population that has had the disease, thus acquiring immunity. Permanent immunity after having the disease is an assumption of the model, but if it happens and how long it lasts is still an open question (see a review of studies in Flodgren 2020).

The simulations of adaptive confinement make it clear the importance of taking early measures against the epidemic. I simulated two scenarios, one with a threshold for confinement of 3500 current infections and another with a threshold of 1000. The first is approximately the number of infected that there were in Spain two days before the confinement was officially declared by the government on the 14^th^ of March, a scenario chosen to represent the actual events. The lower threshold was reached about four days before that, on the 8^th^ of March. The differences between the two scenarios are striking (fig. 4). The peak of the first wave is approximately 108×10^3^ infected with the highest threshold, but only 33×10^3^ with the lower one. More importantly, the model-predicted fatalities reach approximately thirty thousand with the higher threshold (very close to the actual official number) but only *nine thousand* with the lower one; a third of the number of deaths. Thus, the delaying of the start of confinement by a mere four days may have caused a two-thirds increase in fatalities, or twenty thousand more deaths. An earlier confinement would have reduced this number even further. At the start of an epidemic the number of cases rises exponentially, meaning that every single day is important to control the epidemic.

The predictions of the model clearly show that some control strategies and implementations are better that others, even if they are not perfect. They also show that strong measures must be taken very early in the epidemic and very aggressively; that the argument that some measures “cannot be adopted too early” is unjustifiable if the goal is to control the epidemic and save lives. Hopefully, the lessons of modelling and analysis exercises like this and others will be learnt and considered when the next wave arrives and, especially when the next pandemic emerges.

## Data Availability

All data are publicly available. See links below.
Custom code is available at:
https://www.mathworks.com/matlabcentral/fileexchange/77428-fitseir

https://www.ecdc.europa.eu/en/publications-data/download-todays-data-geographic-distribution-covid-19-cases-worldwide

https://github.com/CSSEGISandData/COVID-19

